# AI-driven imputation of a synthetic personality severity index from the NEO-FFI

**DOI:** 10.1101/2025.11.09.25339839

**Authors:** Karin Labek, Roberto Viviani

## Abstract

Personality disorder severity (PDS) is a novel measure of personality in the ICD-11 with potential prognostic value. Due to its recent introduction, however, no information on it exists in most clinical cohorts. Here, we explored the value of its replacement in statistical models by an AI-based synthetic score based on NEO-FFI data, a widely used normative assessment of personality. In linear models with sex, age, traumas in childhood, and occupational status, the synthetic PDS scores reproduced coefficients of the observed scores closely. Unlike the linear model, the neural synthetic scores did not reproduce a spurious association between sex and personality functioning, suggesting that the neural model captured clinically meaningful variance while suppressing confounding demographic signals. Null hypotheses in synthetic models were rejected with only a modest loss of power. These findings suggest that clinically relevant information is latent within normative measures, despite their distinct theoretical origins. The approach enables retrospective severity estimation in datasets that include NEO-FFI items but lack direct assessments of personality functioning, thereby embedding a dimensional, transdiagnostic framework for personality pathology into large health databases without additional data collection and supporting immediate, cross-cohort research on risk and prognosis.

AI-driven imputation of a synthetic personality severity index from the NEO-FFI

## 1. Introduction

In psychiatric diagnosis, sustained critiques of categorical typologies have contributed to a shift toward dimensional transdiagnostic frameworks, as reflected in ICD-11 and DSM-5/DSM-5-TR AMPD (American Psychiatric Association, 2013; World Health Organization, 2022)). This shift has had the greatest impact on the diagnosis of personality disorders. Within this framework, their diagnosis requires grading the severity of impairment in personality functioning across self (identity, self-direction) and interpersonal (empathy, intimacy) domains along a spectrum. Both ICD-11 (non to severe personality disorder) and AMPD (no to extreme impairment) conceptualize this severity dimension analogously, independent of specific trait profiles (Bender et al., 2011; Mulder, 2021; Tyrer et al., 2019). Severity level thus serves as the primary diagnostic domain and represents the core of dimensional personality diagnostic (American Psychiatric Association, 2013; Bender et al., 2011; Sharp & Wall, 2021; Tyrer et al., 2019; World Health Organization, 2022). However, despite its clinical utility and prognostic relevance (Bach & Simonsen, 2021; Hopwood et al., 2011; Mulder & Tyrer, 2019), validated assessments of personality functioning are largely absent from large health and research datasets, as these measures have only recently been introduced. Broad implementation will likely take years, delaying progress toward more precise, individualized diagnosis, prediction, and treatment. Without such measures, opportunities to identify causative factors, understanding mechanism of pathogenesis, and predicting treatment outcomes remain unrealized. In contrast, in the large-scale health databases that have become central resources for mental health research, normative personality measures from the NEO family (e.g., NEO-FFI, (Costa & McCrae, 1992b, 2014)) together with genetic, biological, neuroimaging, behavioural, and lifestyle data are available, providing a basis for deriving proxy indicators of the personality functioning severity in the absence of clinical assessments.

This paper addresses this issue by developing and validating an AI-driven Synthetic Personality Severity Index (SPSI) aligned with ICD-11 personality disorder severity. The SPSI was trained on item-level NEO-FFI responses to enable retrospective estimation of personality-functioning severity when direct assessments are missing. The novelty lies in developing a scalable, trait-based proxy for personality functioning derived from NEO-FFI items—bridging normative and clinical frameworks by harnessing robust prediction in machine learning approaches. The resulting SPSI may be applied to datasets that include NEO-FFI but lack direct personality-functioning assessments, enabling immediate exploratory analyses of associations with established risk and prognostic indicators and supporting cross-cohort, population-scale investigations without additional data collection. In doing so, it leverages existing resources such as large-scale health databases for timely and reliable assessments of personality pathology and promotes sustainability by maximizing the scientific yield of already collected data.

The concept of personality functioning severity marks an important step in personality research (Zimmermann et al., 2020) and aims at providing clinicians with information of prognostic value and about individual treatment needs (Bach & Simonsen, 2021; Buer Christensen et al., 2020; Hopwood et al., 2011; Hutsebaut & Bender, 2024; Mulder & Tyrer, 2019). By contrast, the “Big Five” is rooted in the lexical/factor-analytic tradition of the study of normative trait personality (de Raad & Mlačić, 2017; Wright, 2017). The personality-trait approach offers a complementary but distinct framework for characterizing the broad spectrum of personality variation. However, it was not designed to index clinical severity, even though trait domains can correlate with personality disorders (Costa & McCrae, 1992a; Samuel & Widiger, 2008; Widiger et al., 2017). Therefore, it is of considerable interest to show the extent to which one can be reconstructed from the other, implying that information on personality disorder severity may be present in normative scales. Here, we will show that our model achieves better prediction of personality severity than in its known association with individual NEO-FFI factors. We will also explore the use of this prediction (the SPSI) in imputing personality disorder severity in statistical models to test its association with social and occupational dysfunction, as reported by Hopwood et al. (Hopwood et al., 2011), Morey et al. (Morey et al., 2013) and Buer Christensen et al. (Buer Christensen et al., 2020), and to retrieve its strong links to early trauma.

## 2. Methods

### Ethics information

The study was approved by the Ethical Review Board of the Institute of Psychology of the University of Innsbruck (ethics application No. 79/2021). Participants were recruited via email invitations sent to psychology students at the University of Innsbruck, who were encouraged to share the study with their families and friends. Participants were only included after confirming to have read and agreed to the information about the modalities and finalities of the study.

### Participants and data collection

In this cross-sectional study, 630 participants were initially enrolled in the online survey. The survey was conducted via a web-based application. After identifying and removing duplicates, 591 unique subjects remained in the dataset. We further refined the dataset by excluding participants with session breaks exceeding three hours and those under the age of 18, resulting in 532 subjects. Additionally, three subjects with an average reaction time (RT) of ≤1000 milliseconds were detected using a filtering process based on mean RT per subject. Finally, 43 participants were excluded due to missing data of the questionnaires used in the survey. The final dataset consisted of 486 participants (mean age 24.1, std. dev. 7.57, range 18-80). The gender breakdown was predominantly female (N = 366, 75.31%), with 113 males (23.25%) and seven participants (1.44%) identifying as diverse. Participants reported a range of educational backgrounds: 316 participants (65.02%) had a high school diploma, 108 (22.22%) had a bachelor’s degree, 42 (8.64%) had a master’s degree or diploma, 17 (3.50%) had completed vocational training and three (0.62%) had an intermediate secondary school diploma. In terms of monthly net income, 316 participants (65.02%) reported earning up to €500; 51 participants (10.49%) earned less than €1,000; 45 participants (9.26%) earned less than €1,700; 30 participants (6.17%) earned less than €2,300; 15 participants (3.09%) earned less than €3,000; and 29 participants (5.97%) earned above €3,000.

### Measurements

The training data consisted of the item-level responses from the NEO-FFI (60 items), which assesses the Five-Factor Model of personality—Openness, Conscientiousness, Extraversion, Agreeableness, and Neuroticism (also termed FFM or OCEAN; (Borkenau & Ostendorf, 2008; Costa & McCrae, 1992a, 1992b). ICD-11 personality-functioning severity was assessed with the PDS-ICD-11 self-report (14 items; (Bach et al., 2021); German version: (Zimmermann et al., 2023). In addition to demographics (sex, age, income), we included external validators informed by prior work—social and occupational dysfunction, trauma exposure, and impulsivity—given evidence that personality-functioning severity explains variance in these domains (Buer Christensen et al., 2020; Hopwood et al., 2011; Morey et al., 2013).

#### NEO Five-Factor Inventory

In our study, we used the 60-item German version of the NEO Five-Factor Inventory (NEO-FFI) by (Costa & McCrae, 1989; McCrae & Costa, 2007), adapted by (Borkenau & Ostendorf, 1993) and validated by (Körner et al., 2002). This instrument is the short version of the 240-item NEO Personality Inventory-Revised (Costa & McCrae, 1985, 1989) (NEO-PI-R) and assesses the Big Five personality traits—Neuroticism, Extraversion, Openness to Experience, Agreeableness, and Conscientiousness—using a five-point Likert scale ranging from 0 (“strongly disagree”) to 4 (“strongly agree”). The internal consistencies of the five scales range from α = .72 to α = .87, and the retest reliabilities (over five years) range from r = .71 to r = .82 (Borkenau & Ostendorf, 2008). Internationally, the original 60-item NEO-FFI is the most widely used short version (Körner et al., 2015). The 60-item NEO-FFI in our study demonstrated good internal consistency (Cronbach’s α = .88).

#### Personality Disorder Severity ICD-11 scale (PDS-ICD-11)

The PDS-ICD-11 is a 14-item self-report instrument designed to index personality-disorder severity as specified in ICD-11 (Bach et al., 2021). It covers impairments in self (4 items) and interpersonal functioning (4 items), plus emotion regulation (1), cognition (1), behaviour (3), and overall psychological impairment/distress (1). Items 1–10 are bipolar with five ordered responses (scored 2–1–0–1–2) to capture dysfunction at either pole; items 11–14 are unipolar with four response options (0–3). A summed total yields the severity score. Validation studies indicate a unidimensional latent structure and good convergent validity with established measures (e.g., LPFS-BF 2.0, SASPD, MDPF). The German version, produced via forward–back translation and approved by the original authors, showed comparable psychometrics, including internal consistency of α = .77–.85 and strong convergence with alternative self-reports (Zimmermann et al., 2023). The 14-item PDS-ICD11 scale showed acceptable internal consistency (Cronbach’s α = .79).

#### Adverse childhood experiences questionnaire

Early traumatization was retrospectively assessed with the German version of the ACE questionnaire (Felitti et al., 1998; Schäfer et al., 2009). The ACE is a widely used self-report tool which comprises 10 items regarding abuse (emotional, physical, and sexual), neglect (emotional and physical), separation of a parent, violence against the mother, as well as problems of a household member (substance use, mental disorder, and prison stay). Each item is answered with either yes (1) or no (0), resulting in a sum score between 0 and 10. The German version of the ACE has shown acceptable reliability, with Cronbach’s α = 0.76 (Wingenfeld et al., 2011). In our sample, an acceptable internal consistency of the ACE items could be observed (α = 79).

### Statistical Analysis

Data preprocessing (e.g., exclusion of participants, standardization) and all subsequent statistical analyses were performed in RStudio (version 4.2.1), with the *dplyr* package (Wickham et al., 2023) used for data manipulation. The neural network model was implemented in R via the *keras* package with TensorFlow as computational backend (Chollet & Allaire, 2018) running on Python 3.11.9. Detailed model architecture, training diagnostics, and supplementary statistical results are provided in the Supplementary Material (Appendix) accompanying this article.

### Neural network architecture, training strategy and performance assessment

In a first phase, architecture and hyperparameters of a neural network trained to predict personality functioning scores (PDS-ICD-11) (Bach et al., 2021) from NEO-FFI items (Borkenau & Ostendorf, 2008) were determined and assessed on held-out test sets, i.e. data not used during training. In a second phase, these predicted scores, the ‘neural synthetic personality severity index’ SPSI (*neural* SPSI), were used in linear models with age, sex, occupational status, and early trauma in lieu of the observed scores. For comparison, a second linear synthetic personality severity index (*linear* SPSI) was generated via a linear model with the same personality functioning scores (PDS-ICD-11) as outcome and the five NEO-FFI subscales (Neuroticism, Extraversion, Openness, Agreeableness, Conscientiousness) as predictors. All performance assessments were based on test set data, i.e. data not used in training.

For readers not familiar with neural networks, the *architecture* refers to the way the model is structured. At its simplest, a feedforward network consists of an input layer (here: responses to the NEO-FFI items), an output layer (here: the predicted ICD-11 personality functioning score), and one or more hidden layers in between. Each connection between layers is weighted. During training, the network produces a prediction, compares it to the observed value, and computes an error. This error is then propagated backward through the network (*back-propagation*) to adjust the weights, such that predictions gradually improve with each iteration.

#### Phase 1 – Neural-network training and hyperparameter optimization

The network was trained to predict standardized PDS-ICD-11 scores by back-propagation. The feedforward architecture was deliberately overparameterized to ensure broad model capacity (Belkin et al., 2019) It consisted of six dense hidden layers with rectified linear unit (ReLU) activation (480, 236, 128, 64, 32, and 16 units, respectively), yielding approximately 180,000 trainable parameters. To regularize the model and reduce overfitting, a dropout layer with a rate of 0.40 (40%) was applied after the first dense layer, preventing it from memorizing the training data and thereby improving generalizability. At the top of the chain, a smaller bottleneck layer with eight linear activation units (with L2 regularization with the package default parameter of λ = 0.01) was inserted to compress the learned representation into a lower-dimensional space before passing it to the final output unit predicting the continuous standardized PDS-ICD-11 score (*z*-scores). This architecture encouraged the network to extract efficient, low-dimensional features from the NEO-FFI item responses that were most predictive of personality functioning.

For data partitioning in this phase, the dataset was divided into an exploratory training subset and a test subset. The exploratory subset was used for model fitting and hyperparameter validation, while the test subset was reserved for final evaluation of predictive accuracy. Within the exploratory subset, repeated *k*-fold cross-validation (*k* = 3) was applied. The data were split into three folds (k=3), the model was trained on two folds and validated on the third, repeating the process so that each fold served once as validation. To reduce the influence of a single random partition, the three-fold cross-validation cycle was repeated six times with newly randomized fold partitions, yielding 18 training–validation runs in total. Mean MAE values across epochs (each representing one full pass through the training data, with weights updated after smaller batches of 32 observations) were used to monitor the stability and performance of the chosen hyperparameter configuration (number of epochs, hidden layer sizes, and dropout rate). Based on the cross-validated results of the training data, a training stop at 60 epochs, a batch size of 32, and a linear layer size of 8 units were determined. The final evaluation of predictive accuracy was performed on the held-out test subset.

#### Phase 2 - Evaluation of SPSI use in a linear model and model comparison

After establishing the neural network architecture and training procedure, the model was applied to generate synthetic personality functioning scores (neural SPSI) for use as an imputed variable in a linear model. A bootstrap resampling scheme was implemented: in each of 60 iterations, the dataset was randomly split into 75% training and 25% test data. The network was retrained on the training portion in each bootstrap resample with the hyperparameters established in the previous phase and used to predict synthetic PDS-ICD-11 scores in the test dataset, in which the synthetic scores were compared to the observed scores when used as imputed variables in the linear model.

For comparison, a second linear SPSI set was obtained by fitting a linear model with the same standardized PDS-ICD-11 personality scales as outcome as in the linear network model, and the neuroticism, extraversion, conscientiousness, agreeableness, and openness to experience scales as predictors. The fitted model was applied to the NEO-FFI scales in the test set to compute the comparison SPSI. Note that the neural network was trained from item-level NEO-FFI data, while the comparison SPSI set was based on the scale-level data. The linear model-based SPSI assessed the predictive capacity of a constrained model using previously validated and widely used variables in reporting normative personality traits.

#### Assessment of performance (outcomes)

The performance of the synthetic personality severity index (SPSI) was evaluated for both **neural SPSI** (predicted from item-level NEO-FFI data) and the **linear SPSI** (predicted from domain-level NEO-FFI scales) along two complementary dimensions:

##### Prediction accuracy assessment

As a standard metric of predictive performance, the mean absolute error (MAE) between predicted (synthetic) and observed PDS-ICD11 scores was computed in the test subsets. This procedure enabled us to evaluate both the predictive accuracy of the neural network and the extent to which synthetic scores reproduced the predictive relationships of the observed scores.

##### External validity assessment (association recovery)

The main criterion of success was the accuracy of regression coefficients and *t* test statistics obtained when the SPSI) served as the dependent variable in linear models with established correlates of personality functioning (childhood trauma, log-transformed; age; sex; occupational status). Again, none of these predictors were used during model training. In each bootstrap iteration, SPSI scores were generated in the test subset and regressed on the predefined predictor set. In parallel, an identical model was fit with the observed PDS-ICD-11 score as the outcome on the same resample. The resulting standardized coefficients and *t* statistics were then compared by computing absolute differences between SPSI-based and observed-score estimates, and this similarity was used to quantify how well the SPSI each reproduced external associations. As a complementary check of predictive validity, SPSI was also regressed on the observed PDS-ICD-11 within each test set, and the adjusted R² was recorded.

## 3. Results

### Phase 1: Model architecture and determination of hyperparameters

In the exploratory cross-validation runs (k = 3 repeated six times), the network consistently converged with an average validation MAE of approximately 0.50, indicating stable predictive performance across partitions. When applied to the independent test subset, predictions were strongly associated with observed standardized PDS-ICD-11 scores (r = 0.75). To formally assess predictive accuracy, a linear regression was estimated with observed PDS-ICD-11 scores as the outcome and neural network SPSI as the predictor. Results indicated a strong association (β = 0.95, p < .001), with the model explaining 57% of the variance in observed personality functioning PDS-ICD-11 scores (adjusted R² = 0.57). As illustrated in Figure 2, predicted and observed scores aligned closely along the regression line, with deviations primarily within one standard unit.

**Figure 1.**
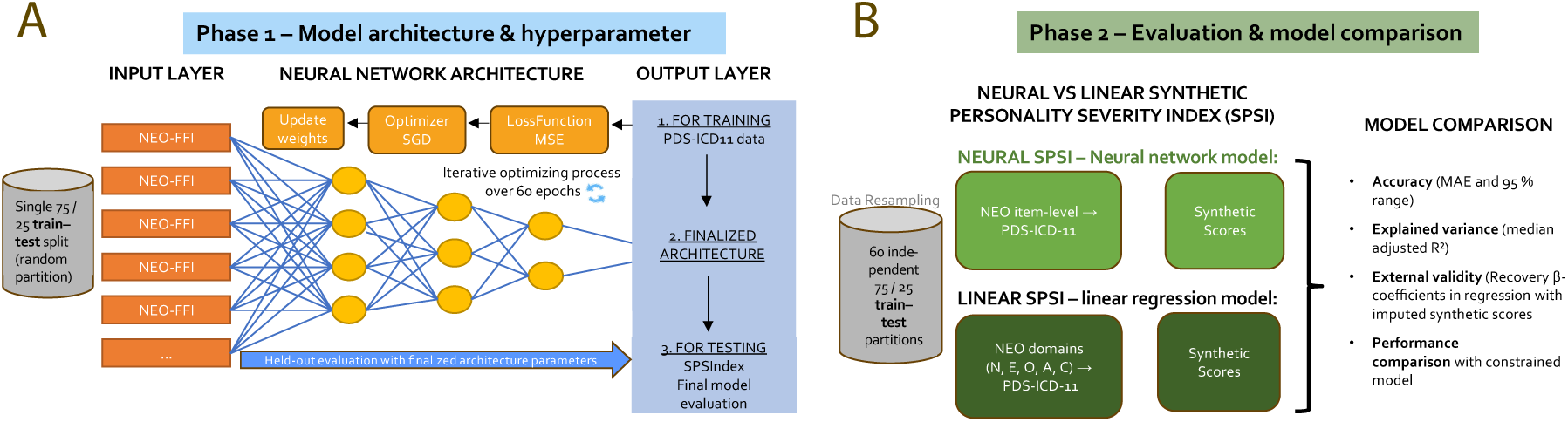
Conceptual overview of model training, validation, and comparison. **(A)** In *Phase 1*, NEO-FFI item-level responses (input layer) were processed through a feed-forward neural network with rectified linear unit (ReLU) activations, optimized via stochastic gradient descent (SGD) and a mean-squared-error (MSE) loss function (orange boxes at the top). The blue box on the right summarizes the three sequential steps of this phase: (1) training the model on 75 % of the data using PDS-ICD-11 scores as targets, (2) finalizing the network architecture and hyperparameters, and (3) testing the model on the held-out 25 % subset to generate the predicted ICD-11 personality-functioning scores (Synthetic Personality Severity Index, SPSI). (**B**) In *Phase 2*, model stability and generalizability were evaluated through 60 bootstrap iterations of independent 75 / 25 train– test splits. The finalized Neural Model (item-level NEO-FFI; light green boxes) and a Linear Model (NEO-FFI domain-level; dark green boxes) were retrained in parallel, each predicting SPSI scores on held-out test data. Across resamples, model performance metrics (MAE, R², β-coefficients) were compared to assess predictive accuracy and external validity, demonstrating robust replication of observed ICD-11 personality-functioning scores (left section of the schematic).

**Figure 2.**
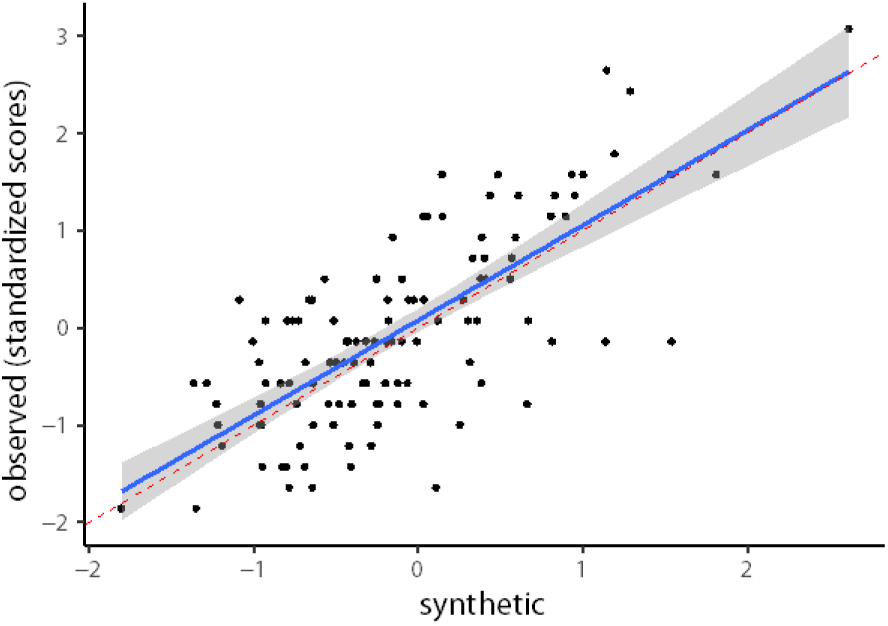
Relationship between observed and synthetic PDS-ICD-11 personality functioning scores. Observed standardized PDS-ICD-11 scores plotted against neural network predictions (neural synthetic SPSI). The regression line (blue) closely follows the 45° identity line (red), indicating good calibration and predictive accuracy.

### Phase 2: Evaluation of SPSI

After determining its hyperparameter in phase 1, the model was retrained and used to generate synthetic personality functioning scores in a resampling scheme (60 iterations with 75% training and 25% test data splits). In the bootstrap sample, the median MAE of the synthetic relative to the observed scores was 0.45, with 95% of all samples lying between 0.42 and 0.47. The adjusted R2 was 0.56, with 95% of the retrained samples lying between 0.47 and 0.66. This confirms that the partition of phase 1 of the study was representative of the data as a whole.

Regression analyses with SPSI synthetic scores as the outcome showed that childhood trauma (log-transformed), age, sex, and occupational status replicated the associations found with the observed PDS-ICD-11 scores (Figure 3). In the boxplot of the left panel (Figure 3A), the coefficients of the models fitted on the synthetic and the observed scores are plotted side by side, allowing one to assess the extent to which the synthetic models were replicating the estimates of the coefficients on the observed personality functioning score in the bootstrapped test sets. Stable predictors such as age and trauma showed narrow distributions in both models, while categorical predictors with small effects (occupation) showed greater variability in both. The boxplot of Figure 3B compares *t* test statistics obtained in the resampled models with the SPIS and with the observed scores. One can see that the ranges of *t* values obtained for the predictors in the synthetic and observed models were similar.

**Figure 3.**
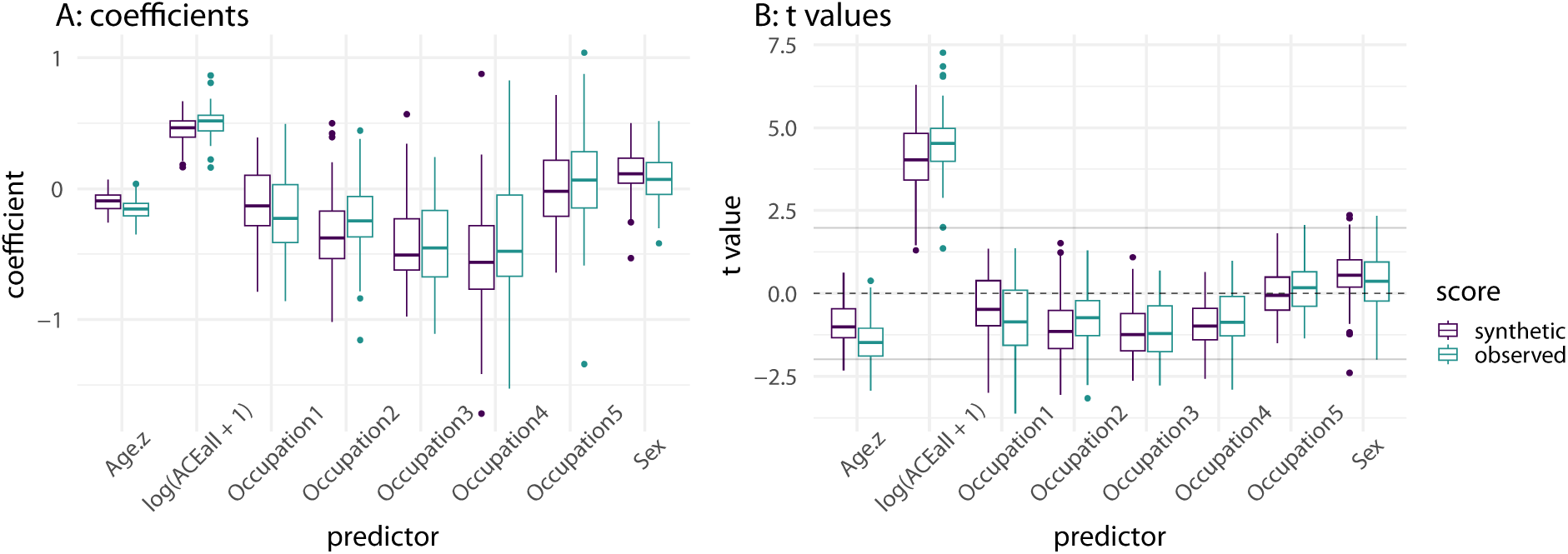
A: comparison of coefficient estimates from bootstrapped samples in model with synthetic and observed PDS scores (neural SPSI). B: comparison of *t* values from the same models. In the case where the null hypothesis holds exactly, *t* values are expected to straddle zero and move beyond the rejection threshold at the false positive bound of the significance level (drawn as two horizonal gray lines for *p* = 0.05, two-tailed, in the figure).

Table 1 provides a more formal assessment of the distributional similarity of coefficients and *t* values in the synthetic and observed models. The first 4 columns concern coefficients and the subsequent 4 columns *t* test statistics. D is the Kolmogorov-Smirnov test statistic for a difference between distributions, which estimates the maximal distance between the two distributions, and p value is the evidence against the null hypothesis of equal distributions. The 50% and 90% columns report the 50th and 90th quantile of the absolute difference between the coefficients or *t* values in the bootstrapped sample. The last three columns report rates of rejection of the null, in percent, in the synthetic model, in the observed model, and their differences.

**Table 1.**
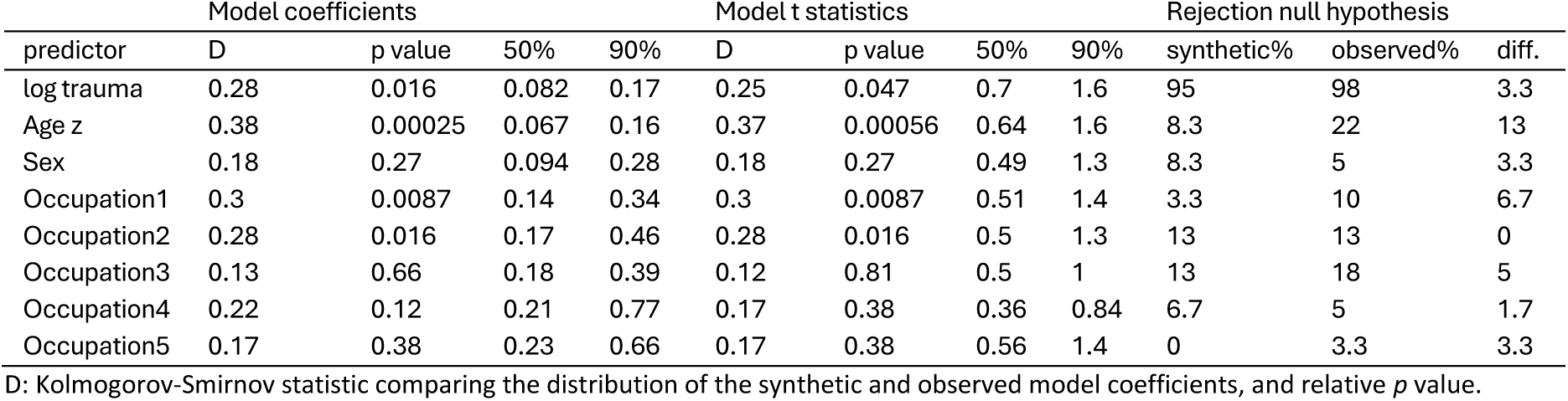
Kolmogorov-Smirnov statistics for coefficients and *t* values, and null rejection rates, neural SPSI

We see here that testing traumas may lead to a difference of 0.70 or less in the *t* test statistics in half of the resamples, and in 1.6 or more in 10% of them. This results in 3.3% differing decisions to reject the null for a sample of this size (N=122). For sex, these values are 0.49 and 1.3, and a similar difference in decision rates. The synthetic model had problems with age, which was underpowered in 13% of tests relative to the observed model.

Together, these results indicate that the synthetic personality functioning score not only achieved robust predictive accuracy but also largely preserved the external correlates of observed ICD-11 personality functioning scores, particularly with trauma and sex, while age was characterized by a modest loss in testing power.

### Assessment of performance (outcomes). Comparison with linear model baseline

To benchmark performance, we also estimated a synthetic personality functioning score using a linear regression model with the five NEO-FFI subscales (neuroticism, extraversion, conscientiousness, agreeableness, openness) as predictors. This more constrained model is based on data in the literature showing that criterion A of the AMPD (the equivalent measure of personality functioning) is associated with neuroticism (Fu et al., 2021; Kotov et al., 2010; McCabe et al., 2021) Negative associations between extraversion and affective symptoms are also reported (Klein et al., 2011; Yang et al., 2024). Table 2 presents the results of a linear regression of the PDS scores on the NEO-FFI subscales in the sample. It shows a strong association between PDS and neuroticism, and to a lesser extent with extraversion, conscientiousness, and agreeableness.

**Table 2.** Regression of PDS scores on NEO-FFI scales

| Predictor | Estimate | Std. Error | t value | Pr(>\|t\|) |
| --- | --- | --- | --- | --- |
| (Intercept) | -0.041 | 0.32 | -0.13 | 0.90 |
| Neuroticism | 0.057 | 0.0037 | 15.64 | < 2e-16 *** |
| Extraversion | -0.013 | 0.0047 | -2.67 | 0.008 ** |
| Openness to experience | 0.0057 | 0.0051 | 1.12 | 0.264 |
| Conscientiousness | -0.017 | 0.0043 | -3.90 | 0.0001 *** |
| Agreeableness | -0.021 | 0.0046 | -4.51 | 8e-06 *** |

Signif. codes: 0 '\*\*\*' 0.001 '\*\*' 0.01 '\*' 0.05 '.' 0.1 ' ' 1. Residual standard error: 0.68 on 480 degrees of freedom. Multiple R-squared: 0.539, Adjusted R-squared: 0.534. F-statistic: 112 on 5 and 480 DF, p-value: <2e-16

As expected from prior reports that linear models can approximate the performance of more complex machine-learning approaches (Hastie et al., 2009), this model achieved considerable levels of accuracy. However, the MAEs of the linear model-based SPIS were consistently higher than the neural network SPSI (median 0.53, compared to 0.47 in the neural network, with 95% of samples lying between 0.51 and 0.55, showing no overlap with the analogous interval in the neural network). The median adjusted R2 was 0.52 (instead of 0.56) with 95% of the samples lying between 0.41 and 0.62 (the neural network was between 0.47 and 0.66).

Furthermore, across all predictors of interest (trauma, age, sex, occupation), the accuracy of coefficients was equal or inferior to that of the feedforward neural network. Accuracy gains for occupation were smaller and more variable, reflecting its lower true association with personality functioning (Figure 4). Overall, the feedforward neural network not only improved predictive accuracy but also provided more reliable estimates of external associations than a linear regression baseline.

**Figure 4.**
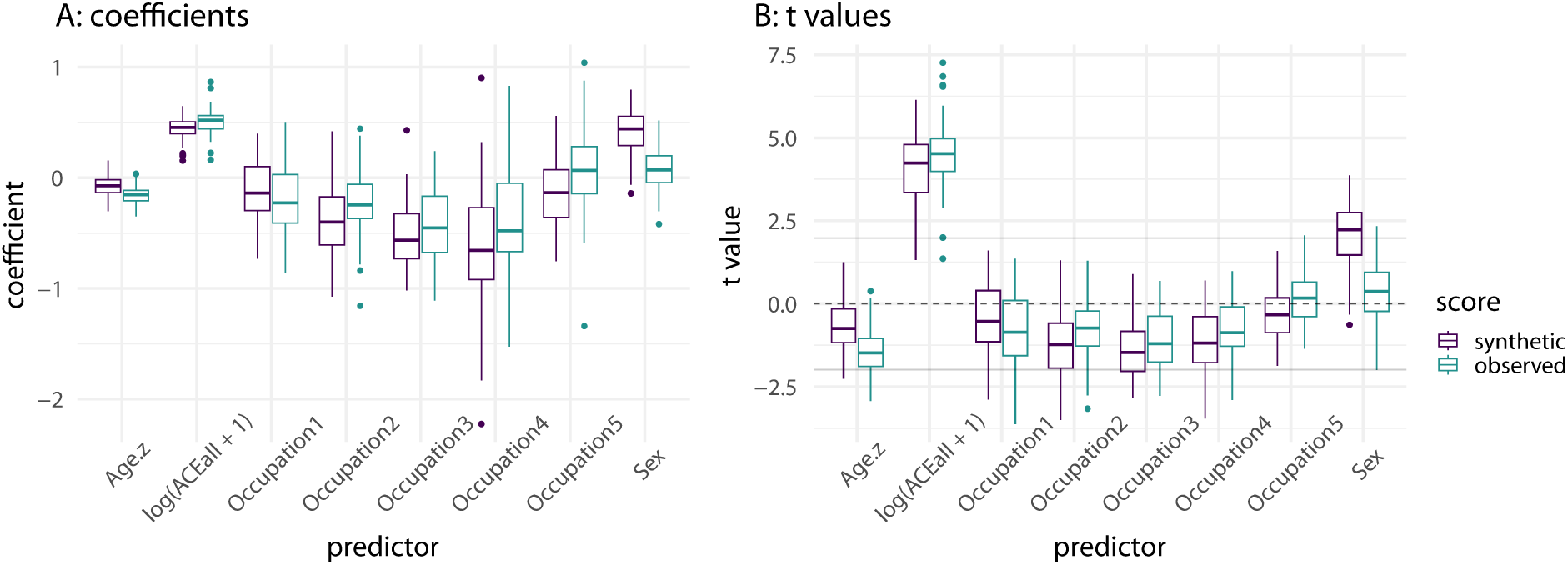
Comparison of coefficients (A) and *t* values (B) in the synthetic and observed models with synthetic PDS scores obtained from a linear model of PDS from NEO-FFI scales (linear SPSI).

Table 3 presents the Kolmogorov-Smirnov statistics of the distributional divergence between linear model-based synthetic and observed models. The D statistics was always higher than in the AI-based model, and the differences in null rejection rates much higher.

**Table 3.**
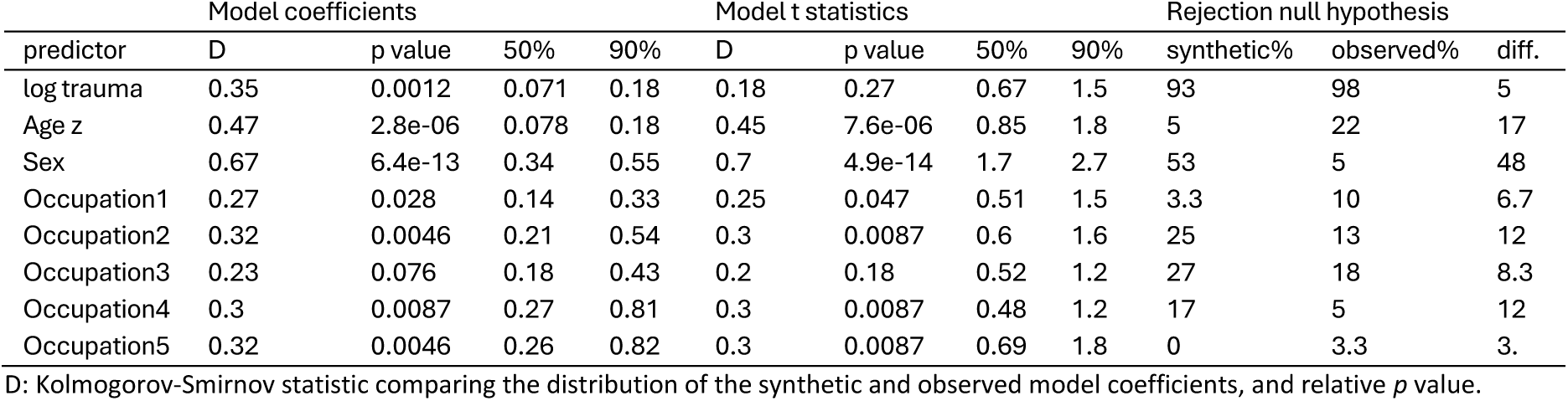
Kolmogorov-Smirnov statistics for coefficients and *t* values, and null rejection rates, linear SPSI

The neural network-based and the linear regression-based SPSI performed very differently when used to assess the association of personality functioning with sex. As is apparent in the Figure, *t* tests statistics values for sex were much higher in the linear regression-based SPSI, indeed much higher than the *t* values obtained from the observed personality functioning scores. As a consequence, the null hypothesis of an association between PDS and sex was rejected in only 5% of the resampled models with the observed score, but in 53% of the synthetic models. This appears to be due to the linear model biasing the predicted SPIS towards associations that are particularly well represented in the NEO-FFI scales, but not in personality functioning. A logistic regression to predict sex from the observed personality functioning scores gave a non-significant result (*z* = 0.41, *p* = 0.16), indicating that observed personality functioning scores were only weakly or not associated with sex. In contrast, in these same data, neuroticism was a strong predictor of sex (*z* = 6.21, *p* < 0.001). Even if the neuroticism items are seen by the neural network model, its synthetic scores did not show the tendency to associate with sex seen in the linear model.

In conclusion, the linear model-based SPSI were slightly less accurate than those from the neural network model. This moderate difference, however, was magnified when these SPSI scores were used in the linear model, where they tended to reproduce associations present with the scales instead of those of the observed personality functioning scores. In particular, the neural network-based SPSI avoided the spurious sexual association bias of the NEO-FFI transmitted to the linear SPSI.

## 4. Discussion

The present study demonstrates the feasibility of deriving a synthetic personality severity index (SPSI) that approximates ICD-11 personality functioning (Bach et al., 2021) from NEO-FFI (Borkenau & Ostendorf, 2008; Costa & McCrae, 2014) item-level data using machine learning. This approach addresses a critical gap in psychiatric and psychological research: while personality functioning has been recognized as the central dimension of personality disorder diagnosis in ICD-11 and DSM-5-AMPD (American Psychiatric Association, 2013; Bender et al., 2011; Sharp & Wall, 2021; Tyrer et al., 2019; World Health Organization, 2022), validated measures of this construct remain largely absent from large-scale health datasets. In contrast, normative personality data such as the NEO-FFI (Costa & McCrae, 2014) are widely available in cohort studies, biobanks, and genetic consortia, often alongside rich biological and clinical information. Our findings indicate that these datasets may already contain sufficient information to approximate personality functioning severity, thereby opening avenues for retrospective and large-scale analyses that were previously impossible. Beyond methodological advances, the SPSI illustrates how machine learning can help to conserve resources by unlocking the latent clinical utility of existing large-scale datasets, thereby advancing research in personality pathology until new, dedicated data are collected.

Personality functioning represents a major advancement in psychiatric nosology, addressing the limitations of earlier categorical systems by capturing severity as a continuous, transdiagnostic construct (Morey et al., 2022; Sharp & Wall, 2021; Zimmermann et al., 2019). It is increasingly recognized as a robust predictor of prognosis, therapy outcome, and treatment needs (Bach & Simonsen, 2021; Hopwood et al., 2011; Mulder & Tyrer, 2019), with implications across a broad range of disorders (Ma et al., 2024; Zimmermann et al., 2019). Moreover, personality functioning plays a pivotal role in the establishment of the therapeutic relationship, a prerequisite for successful psychotherapy and medication adherence (Bach & Simonsen, 2021). Being able to measure this dimension reliably, even retrospectively, thus carries substantial clinical and public health relevance.

This study bridges two independently developed frameworks: the lexical/factor-analytic tradition of normative personality research (Big Five) (de Raad & Mlačić, 2017; Wright, 2017) and the clinically anchored ICD-11 concept of personality functioning severity (Bach et al., 2021; Tyrer et al., 2019). Using neural networks, we showed that a clinically relevant signal can be extracted from NEO-FFI item data to approximate ICD-11 severity, even though the NEO-FFI instrument was not designed for this purpose. The SPSI may provide a methodological bridge between non-clinical personality frameworks and clinical psychiatry, demonstrating the feasibility of mapping normative traits onto clinically meaningful measures of personality functioning. Because the SPSI is trained at the item level, it may preserve fine-grained signal that is typically lost when only domain scores are used. Compared to a linear baseline, the neural network improved both prediction accuracy and recovery of external associations, highlighting its value for modeling non-linear and interactive item relations.

A particularly noteworthy finding was that the neural network-based SPSI did not reproduce the spurious association between sex and personality functioning that emerged in the linear regression-based model. This suggests that the neural model learned to disentangle the clinically meaningful variance related to personality functioning from variance associated with demographic or trait-related biases present in the NEO-FFI. In other words, the nonlinear mapping allowed the network to *“see through”* surface correlations among NEO-FFI scales and to extract latent information patterns more specifically predictive of functioning severity. This property is consistent with the capacity of overparameterized deep models to capture complex interaction structures and to suppress redundant or confounded signals when trained with appropriate regularization (Belkin et al., 2019).

Conceptually, this implies that the neural SPSI may not merely reproduce the profile of the Big Five traits but reconstructs a distinct dimension that aligns more closely with the clinical construct of personality functioning as defined in PDS-ICD-11 (Bach et al., 2021; Tyrer et al., 2019), as highlighted by the differing associations with sex. The fact that neuroticism, strongly linked to both sex and general distress, was visible to the network but did not inflate sex associations in the synthetic scores indicates that the model effectively adjusted for the shared variance between neuroticism and sex while retaining the variance truly related to functioning. This capacity can be viewed as an emergent form of “implicit deconfounding,” where the network isolates the target construct from correlated but non-causal inputs.

The SPSI also supports emerging directions in mental health informatics, including (a) personalized and stratified care, by enabling severity-based cohort stratification and outcome prediction; (b) preventive approaches, by allowing early identification of individuals at elevated impairment risk in population cohorts, and (c) computational “digital twin” frameworks (Moggia et al., 2024; Spitzer et al., 2023), by supplying a scalable, continuously updatable severity signal that can be integrated with longitudinal behavioural, biological, or service-use data to simulate trajectories and intervention effects. These developments have accelerated with wider availability of large, linked datasets and advances in model training and deployment.

In sum, repurposing normative trait data with modern machine-learning methods provides a practical, clinically anchored route to embed dimensional personality diagnostics in existing resources. This can accelerate discovery, support precision-medicine initiatives, and enhance the sustainability of population-scale mental-health research.

## Data Availability

All data produced in the present study are available upon reasonable request to the authors and after verifying that the purposes of any new use are compatible with those stated in the original consent given by participants

## Limitations

Several limitations warrant consideration. First, the SPSI is an empirically derived proxy and should not be interpreted as interchangeable with clinician-rated interviews; clinical decisions should continue to rely on direct assessment where feasible. Second, the study was based on a single, moderately sized dataset, and categorical predictors with small effects (e.g., occupational status) showed greater variability, underscoring the need for replication in larger and more diverse cohorts. Third, residual confounding may influence recovered associations; pre-registered analyses and sensitivity checks are recommended. Fourth, the current model is trained on cross-sectional data; prospective validation against longitudinal outcomes (e.g., treatment response, relapse, functional change) is needed.

## Supplementary material

In this supplementary text, we present the results and the technical justification for the development and evaluation of a feedforward neural network to predict ICD-11 personality functioning (assessed with the PDSICD11 scale) from normative NEO-FFI data. The predicted ICD-11 personality functioning score is used as a synthetic imputed score in lieu of the original score and is evaluated by using it as the outcome in a linear model with sex, age, trauma in childhood, and occupational status as predictors.

### S1. Specification of the model

We trained here a densely connected feed-forward neural network with rectified linear units to predict personality functioning from responses to NEO-FFI individual items. Back-propagation was used to compute the gradient of the error function, which was here mean square error. Training was conducted with stochastic gradient descent. The personality functioning scores were standardized (within the training and test samples separately) to aid interpretability of mean absolute errors (MAE) as an index of accuracy.

To provide a very wide model space and let the network itself identify the model appropriate model, we used a large number of weights, with a drop-out layer of 40% after the first layer. Such overparametrized networks have been shown to perform better than smaller architectures and to be less prone to overfitting (see Glorot et al. 2011, Belkin et al. 2019; see also Hastie et al., 2022). At the top of the chain, we inserted a smaller layer with eight linear activation units to factorize the output prior to forming the prediction of the personality functioning score. With a larger dataset, it might be possible to explore performance changes depending on the size of this layer; here, we found that eight units gave performance comparable to small deviations from this size.

The model was specified in R using the bindings to the *keras* package, with TensorFlow as the computational engine using Python 3.11 endpoints (Chollet & Allaire, Deep Learning with R, Manning, 2018).

### S2. Hyperparameter tuning and training diagnostics

To determine the net architecture (including the size of the last linear layer), we preliminary divided the data into a training and test set. Within the training dataset, the model was evaluated with K-fold cross-validation with 3 folds. We recycled these folds to get a more accurate assessment of the performance of the model.

**Figure S1.**
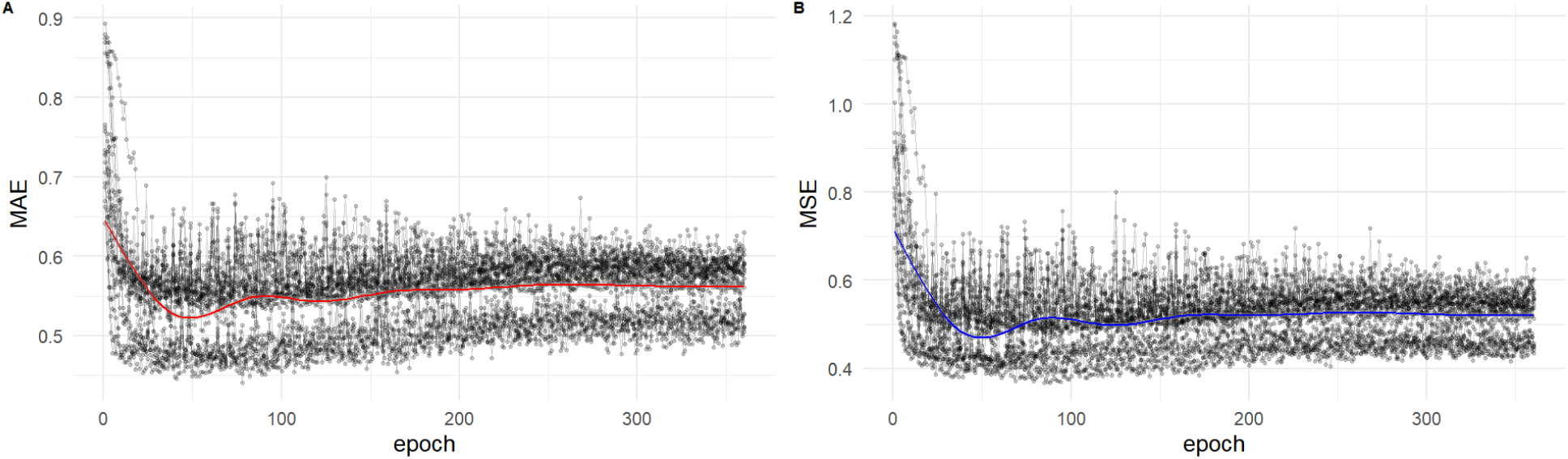
Training diagnostics of the overparameterized neural network model. Panel A displays the mean absolute error (MAE), and Panel B the mean squared error (MSE) over 360 training epochs, each based on a resampled training set. Each grey dot represents the loss at a given epoch from a bootstrap-resampled training run, with the smoothed trend shown in red (A) and blue (B).

The overparametrized learning architecture showed fairly robust and constant performance (as assessed with the mean absolute error relative to the target). Based on the resampled training set, we selected a batch size of 32 and a stopping of training at epoch 60. This resampled training set was also the basis of other modelling choices mentioned earlier, such as the size of the top linear layer.

Note, however, that this overparametrized architecture did not overfit much. We could verify that these parameter choices had a minor effect only on the outcome; see Belkin et al. 2019). For comparison, the average MAE at the epoch 60 was 0.54, while at the final epoch 360 considered in the exploratory training, it was 0.57.

### S3. Predictive Performance (neural vs linear Models)

We then proceeded to train the model on the whole training set and test its performance on the test set by regressing the observed standardized scores on the synthetic scores.

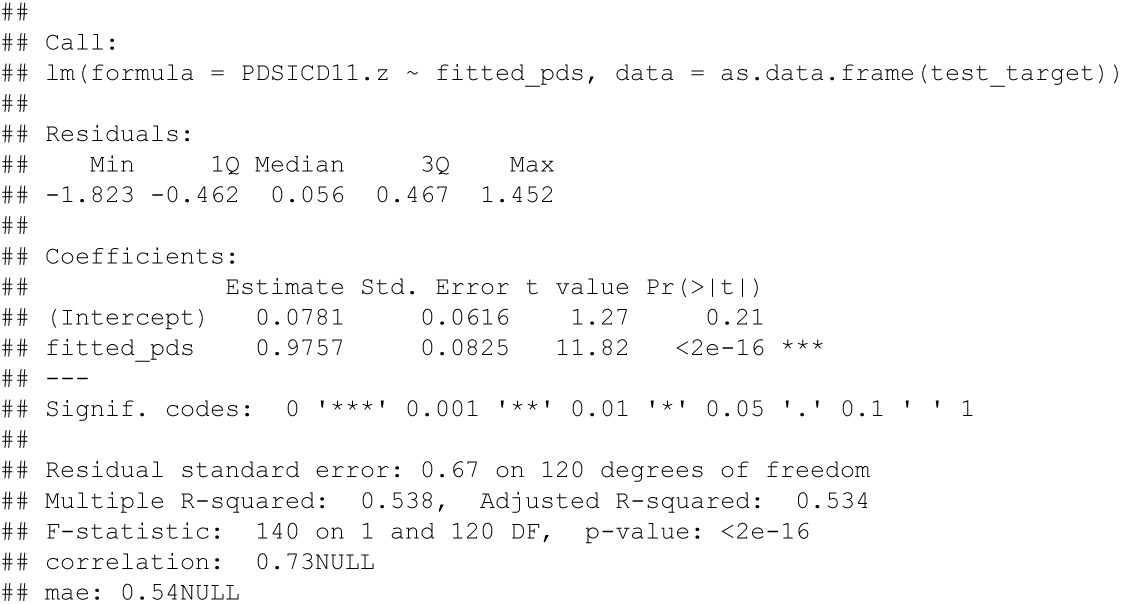

The MAE was 0.54, indicating that most synthetic indices were below one standard deviation of the observed scores (the scores were standardized). The regression of the observed scores on the synthetic scores gave an adjusted R^2^ of 0.53, an estimate of the shared variance. The regression also produced a coefficient slightly less than unity, however with a confidence interval that included unity. We can visualize all this in the scatterplot shown above. The confidence intervals of the fit (blue line and grey area) include the 45° diagonal line from the origin (red line), and most points (the observed scores) are scattered within one standard deviation of the fit (the synthetic score).

**Figure S2.**
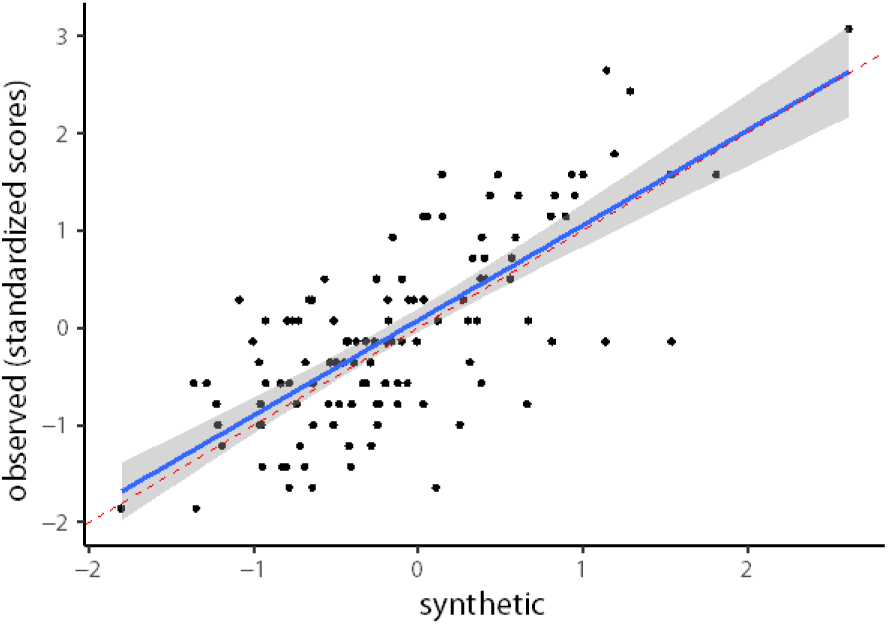
Relationship between observed and synthetic PDS-ICD-11 personality functioning scores. Observed standardized PDS-ICD-11 scores plotted against neural network predictions (neural synthetic SPSI). The regression line (blue) closely follows the 45° identity line (red), indicating good calibration and predictive accuracy.

These results may be interpreted as follows. The slope near unity means that a difference in the predicted score of one standard deviation corresponds to about the same difference in the observed scores. For this reason, the line of the fit is almost identical to the 45° diagonal line going through the origin, which represents the case where predicted and observed scores are identical.

### S4. Association recovery and bootstrap analyses

A more comprehensive assessment of the outcome may be given by resampling these partitions in a bootstrap scheme, providing information on the variation that may be expected in the bias and accuracy of the synthetic scores. We therefore resampled the partition 60 times, producing as many samples of synthetic and observed personality functioning scores in the test sets (when forming the partition, the datapoints are resampled without replacement to avoid information leaks between the training and the test sets, but the partition themselves are sampled without replacement).

The median MAE of the bootstrap resampling was 0.45, with 95% of all samples lying between 0.42 and 0.47. The adjusted R2 was 0.56, with 95% of the retrained samples lying between 0.47 and 0.66.

We now address the issue of the use of the synthetic score in a statistical model, comparing models with the synthetic and with the observed personality scores. The same 60 training and evaluation bootstrap resampling sets of the previous section were used. Personality functioning (synthetic or observed) was the outcome variable, while sex, age, traumas in childhood, and education level were predictors. Importantly, the neural network that gave the synthetic scores was trained without using these predictors of personality functioning.

**Figure S3.**
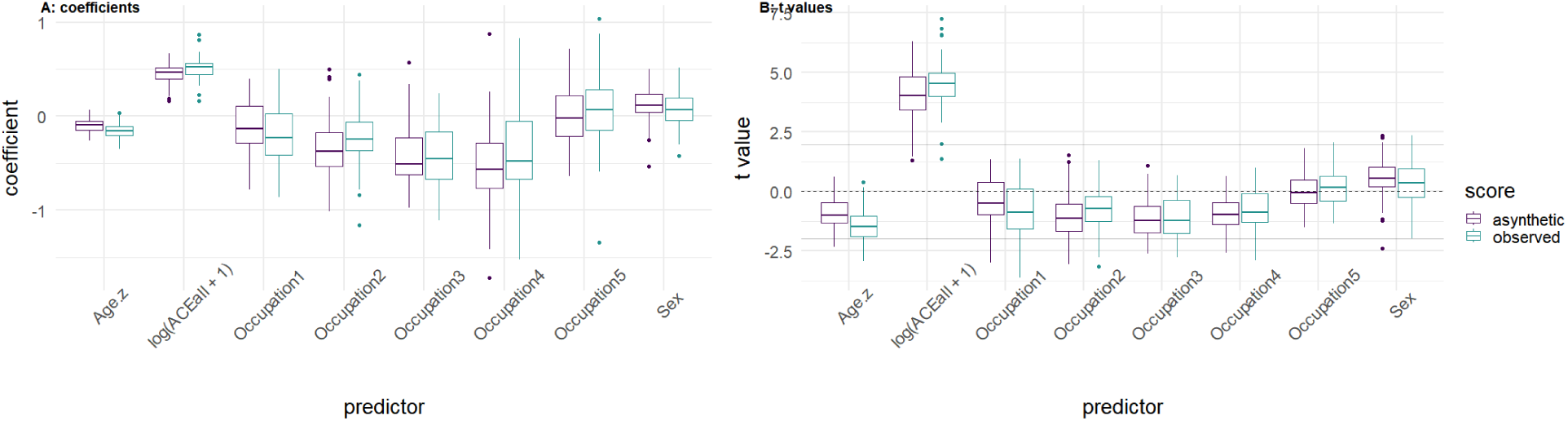
neural network-based synthetic score evaluation. A: comparison of coefficient estimates from bootstrapped samples in model with synthetic and observed PDS scores (neural SPSI). B: comparison of *t* values from the same models. In the case where the null hypothesis holds exactly, *t* values are expected to straddle zero and move beyond the rejection threshold at the false positive bound of the significance level (drawn as two horizonal gray lines for *p* = 0.05, two-tailed, in the figure).

In the boxplot of panel A of the figure below, the coefficients of the models fitted on the synthetic and the observed scores are plotted side by side. Because the bootstrap gives an estimate of the distribution of the resampled statistic, this boxplot allows one to assess the extent to which the synthetic models were replicating the distribution of the estimated coefficients on the observed personality functioning score in the test sets.

In panel B, the t test statistic values are plotted. Recall that, if the null is true, the dispersion in the distribution of a t value only depends on its degrees of freedom and spans values across zero. The plot shows the two-tailed 5% rejection thresholds, which should be crossed by about 5% of t values if the null is true.

The following table provides a more formal assessment of the distributional similarity of coefficients and t values in the synthetic and observed models. the first 4 columns concern coefficients and the last 7 the t test statistics. The statistic column provides the Kolmogorov-Smirnov D test statistic for a difference between distributions, which estimates the maximal distance between the two distributions, and *p* value is the evidence against the null hypothesis of equal distributions. The 50% and 90% columns report the 50th and 90th quantile of the absolute difference between the coefficients or t values in the bootstrapped sample. The last three columns report rates of rejection of the null, in percent, in the synthetic model, in the observed model, and the difference between these two.

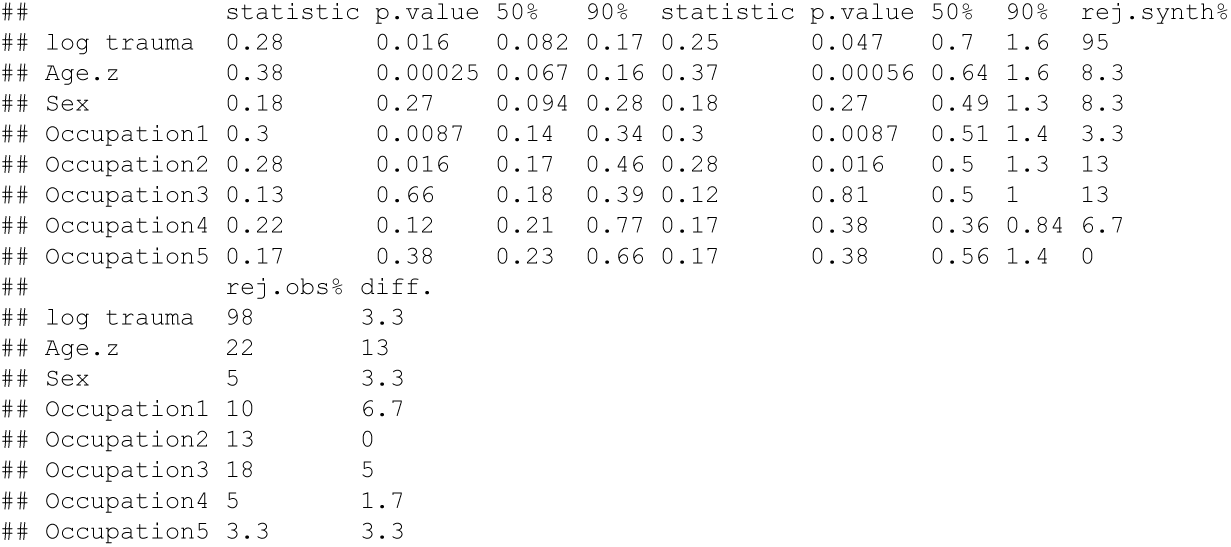

We see here that testing traumas may lead to a difference of 0.70 or less in the t test statistics in half of the resamples, and in 1.6 or more in 10% of them. This results in 3.3% differing decisions to reject the null for a sample of this size (N=122). For sex, these values are 0.49 and 1.3, and a similar difference in decision rates. The synthetic model has problems with age, which is underpowered in 13% of tests relative to the observed model.

### S5. Comparison of bootstrapped coefficients with full-sample estimates

The following box-plot compares the coefficients obtained from the bootstrapped regressions of the predictors on the synthetic personality score with the coefficients obtained from the regression on the observed scores on the whole sample (red points, no resampling). One can see that the coefficients of the observed models are closed to the medians of the bootstrapped coefficients of the synthetic models, and were always falling within the interquartile ranges.

**Figure S4.**
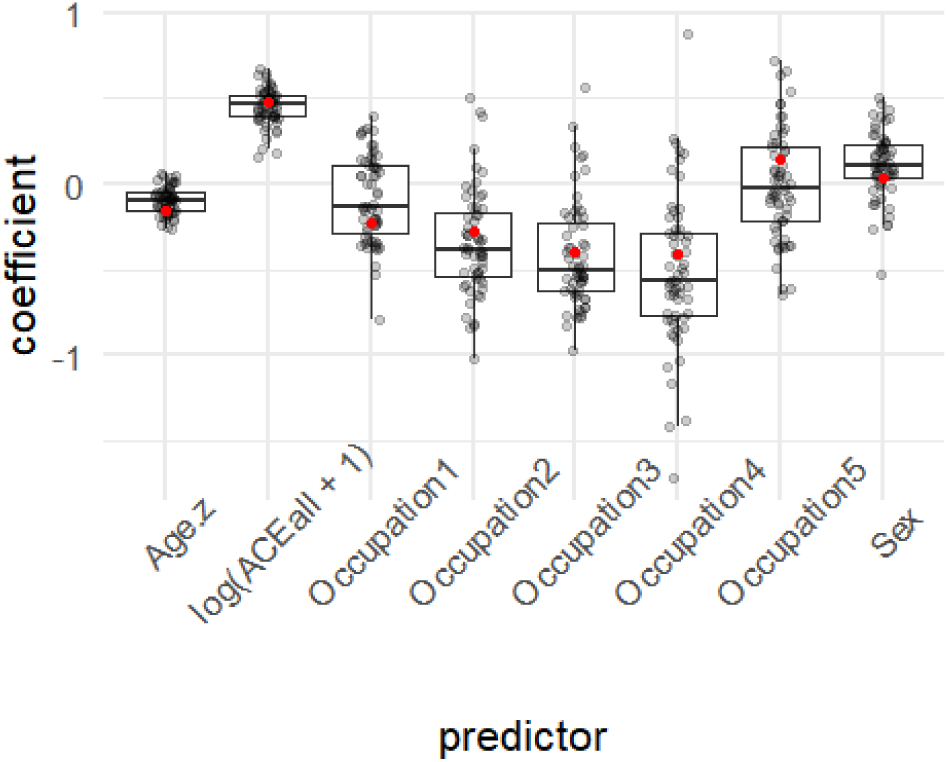
Boxplot of bootstrap resampled regression coefficients for synthetic personality functioning scores (neural network-based) compared to coefficients from the observed scores. The figure shows the distribution of regression coefficients from 60 bootstrap samples in which the synthetic personality functioning scores (predicted by the neural network) were regressed on a common set of predictors: standardized age (Age z), log-transformed adverse childhood experiences (log(ACEall + 1)), occupational status (dummy-coded as Occupation 1–5), and sex. Black dots represent individual bootstrap sample estimates, boxplots reflect their interquartile ranges, and red dots indicate the corresponding coefficients obtained from the regression using observed PDS-ICD-11 scores in the full dataset (without resampling).

### S6. Performance comparison of linear and neural network-based synthetic personality functioning models

It has been recently noted that criterion A of the AMPD is associated with neuroticism (McCabe, Oltmanns, & Widiger 2021, Kerber et al. 2024). Here, we are predicting personality functioning as assessed by the ICD-11. However, these two indices of personality functioning are closely related, so we may expect several association between the NEO-FFI and the personality functioning score used here. The model below shows the associations found empirically in our dataset.

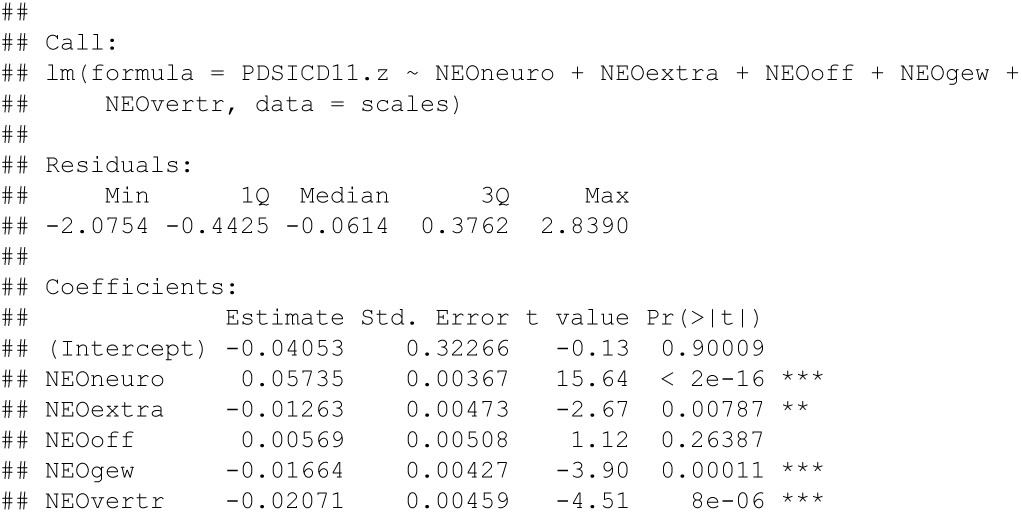
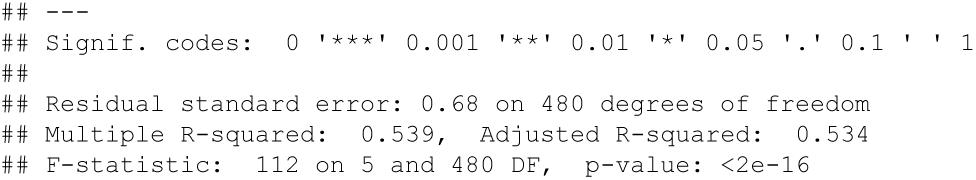

Similarly to what reported in the literature for criterion A of the AMPD, neuroticism was a strong predictor of personality functioning. Also agreeableness was negatively associated with it, and the association with conscientiousness should not surprise given its known association with self-regulation.

We composed a synthetic personality functioning score from the five subscales of the NEO-FFI using this linear model and the same setup as in the previous section. Our objective was to verify that the neural network that we trained performs better than this simple predictor of personality functioning.

**Figure S5.**
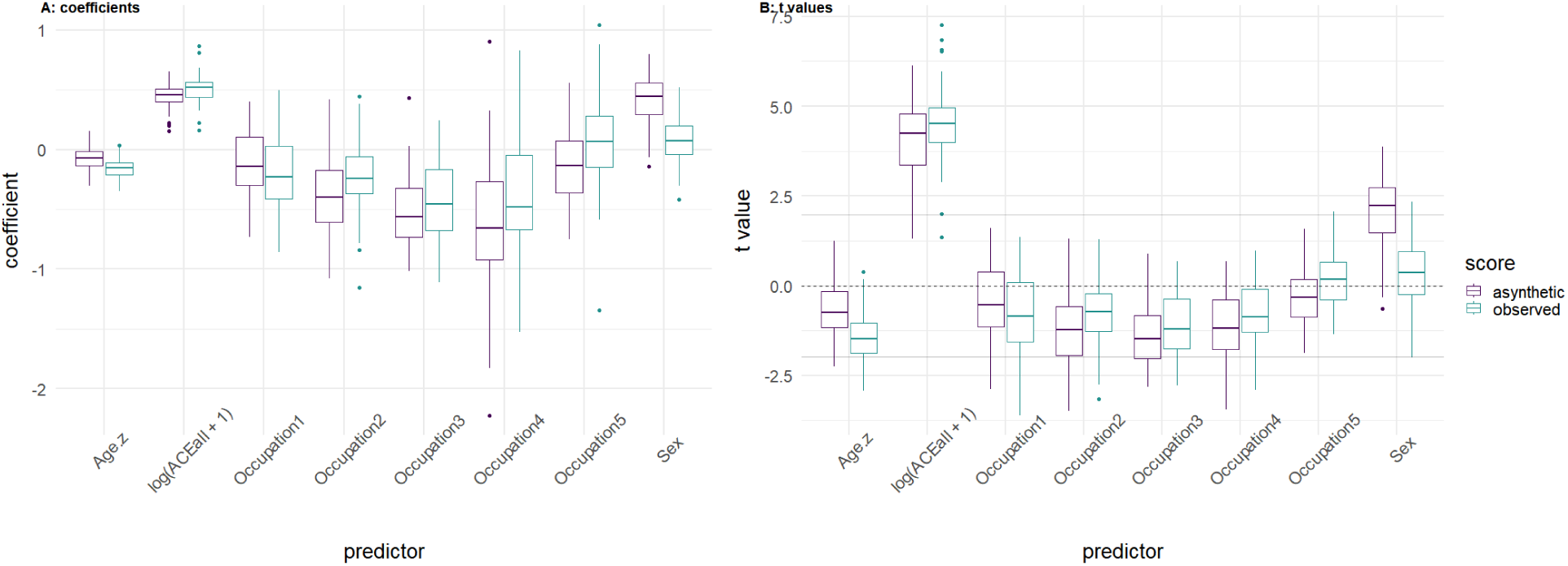
linear model-based synthetic Score evaluation. Comparison of coefficients (A) and *t* values (B) in the synthetic and observed models with synthetic PDS scores obtained from a linear model of PDS from NEO-FFI scales (linear SPSI).

The MAE of the resampled NEO models was 0.53 (compared to the 0.45 of the neural network), and 95% of the resampled MAE were comprised between 0.51 and 0.55 (with no overlap to the analogous interval for the resampled neural networks). The median adjusted R2 was 0.52, with 95% intervals between 0.41 and 0.62 (the neural network was between 0.47 and 0.66).

Below, we draw the boxplots of the resampled coefficients and t values of the synthetic and observed models.

The coefficients of the linear model-based synthetic model are further away from those obtained with the observed score than in the synthetic model from the neural network. To compare the two, we plotted below the mean absolute errors of the coefficients, relative to those obtained in the model with the observed scores.

**Figure S6.**
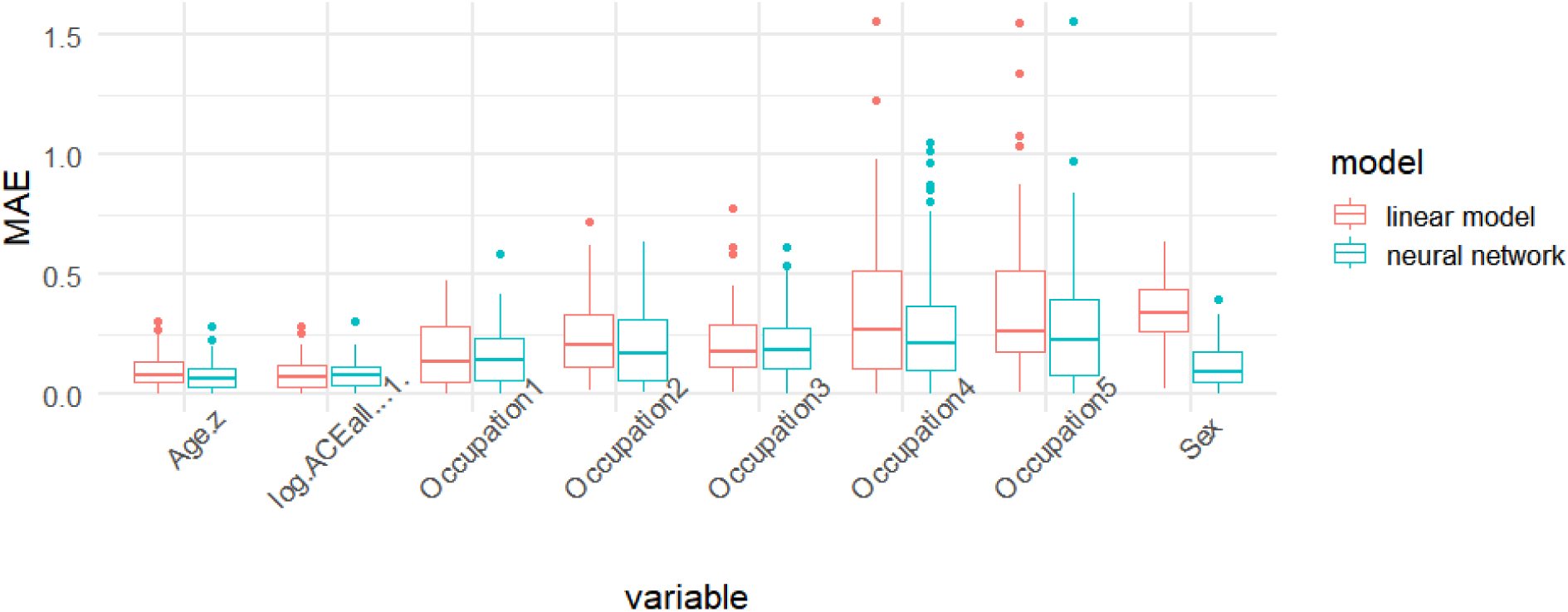
Mean Absolute Error (MAE) comparison between neural network– and linear model–based synthetic personality functioning models. Distribution of MAE of regression coefficients across 60 bootstrap iterations for each predictor variable. Blue boxes represent the neural network–based synthetic personality severity index (SPSI) derived from item-level NEO-FFI data, while red boxes represent the linear model–based SPSI derived from NEO-FFI domain scores.

The neural network model always outperformed the linear model except for traumas and occupation1, where the performance was similar.

The inferior performance of the linear model-based synthetic scores is confirmed in the assessment of the distributions of coefficients and t values.

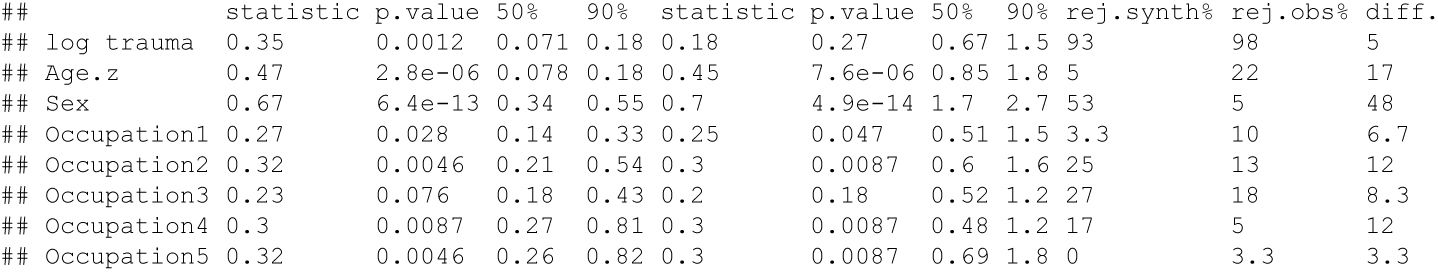

One can see that the divergence in the distribution of estimates in the two models is equal or higher than in the comparison with the neural network-based synthetic scores, leading to different rejections of the null except for occupation5. This divergence is particularly severe for sex.

### S8. Qualitative evaluation of trait confounding and sex association

The synthetic model does a much better job in predicting the observed associations of the personality scores to age and sex. This is because the linear model for the synthetic scores may bias the prediction towards associations that are particularly well represented in the NEO-FFI summary scales. For example, a logistic regression to predict sex from the observed personality functioning gave a non-significant result:

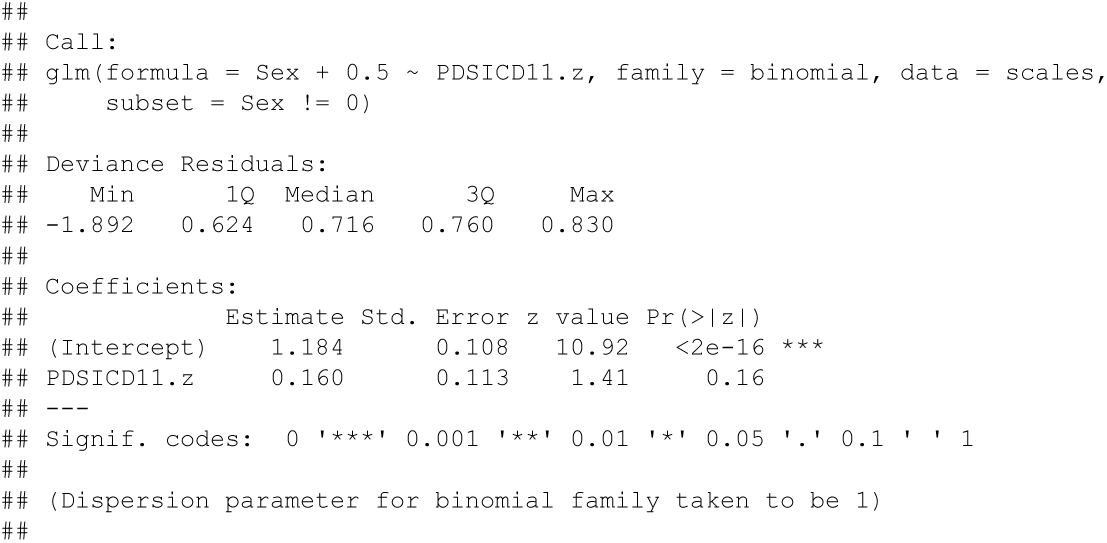
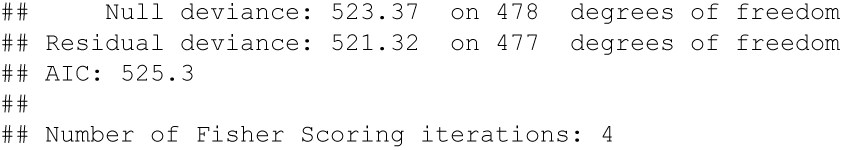

In contrast, in these same data neuroticism was a very strong predictor of sex:

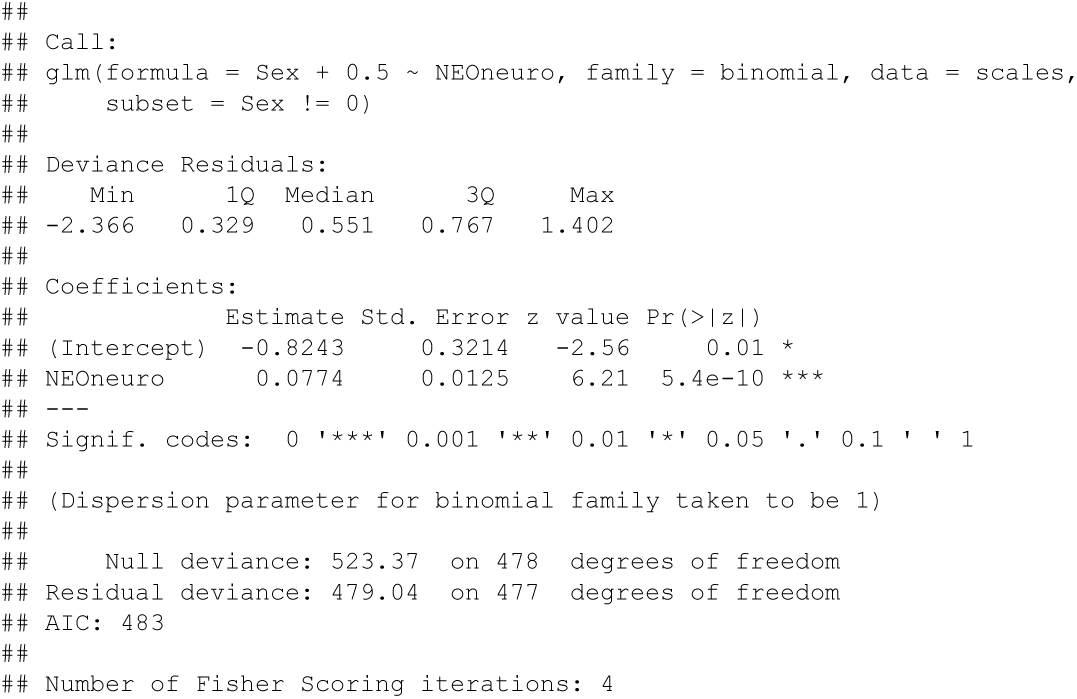

As shown just above, the range of t values produced in the bootstrap resampling by the model using the neural network synthetic personality score overlapped with those produced by the the model with the observed scores. In conclusion, the neural network avoided some spurious associations that were present in the linear prediction of personality scores from the NEO-FFI.

